# Development of a Machine Learning Tool for Home-Based Assessment of Periodontitis

**DOI:** 10.1101/2025.03.10.25323689

**Authors:** Zoe Xiaofang Zhu, Xingwen Wu, Lifang Zhu, Naciye Uzel, Athanasios Zavras, Qisheng Tu, Jake Chen

## Abstract

According to an ADA report, approximately 15% of the US population requires dental care annually but does not receive it. Access to dental care, particularly for periodontal examinations, is challenging for many individuals, leading to uncontrolled periodontitis progression and systemic health complications. Periodontitis, an inflammatory gum disease, affects nearly half of American adults over 30. Current diagnostic approaches rely on periodontal exams and radiographs, requiring clinical settings and experienced dental care providers. However, many individuals lack access to dental care, making it difficult to obtain up-to-date clinical probing depth, dental X-rays or CT scans.

To address this gap, we developed a machine learning (ML) tool for at-home preliminary periodontitis assessments. This tool would benefit individuals unaware of their undiagnosed periodontal conditions and those with limited access to dental care, empowering them to prioritize dental care and seek timely treatment within their constraints.

Our tool leverages the NHANES database to train an ML model on multimodal features relevant to periodontitis that are radiographic-independent. We labeled the individuals with different periodontitis severity based on their periodontal charting records and performed feature engineering on the dataset. We first developed a baseline model and subsequently trained additional classifiers, conducting a comprehensive hyperparameter search that resulted in consistent performance. The best-performing model was evaluated on the test set, achieving an overall precision of 0.80 and AUC of 0.81, demonstrating robust classification performance without overfitting. Feature importance analysis provided guidance for the questionnaire design for the real-world application of this tool. Additionally, our novel approach of analyzing misclassified populations offered insights for data interpretation, supported model improvement, and revealed deeper correlations between periodontitis and its risk factors.

Our model exemplifies the capacity to leverage extensive public health databases for periodontitis evaluations. Ultimately, our ML-driven tool aims to overcome existing dental care barriers by providing users with periodontitis predictions and personalized dental care suggestions, all easily accessible from their smartphones or laptops at home.

**One sentence summary:** In this study, we developed the machine-learning (ML) model for home-based screening of periodontitis and demonstrated that non-radiographic data—encompassing demographic and nutrition information, as well as medical and oral health conditions—possesses strong predictive power for periodontitis, empowering individuals with limited access to dental care to properly identify their periodontal health status and promote timely intervention.

## 1. Introduction

Periodontitis, a prevalent form of gum disease and a common oral health concern, can have a profound impact on an individual’s well-being. “It can lead to chronic inflammation, bone loss, and ultimately, tooth loss (Abusleme et al. 2021). Moreover, periodontitis is also linked to various chronic conditions, including diabetes, atherosclerosis, stroke, and Alzheimer’s disease (Sim et al. 2008; Chapple et al. 2013; Dietrich et al. 2013; Liccardo et al. 2019; Larvin et al. 2023). According to the CDC, nearly half of American adults over 30 suffer from periodontal disease, with the prevalence rising to 70% among those aged 65 and older (Eke et al. 2020).

While treatment options such as professional cleaning, antibiotics, scaling and surgical interventions are available, making periodontal disease usually controllable and treatable, limited accessibility to dental care presents a formidable impediment for individuals facing economic constraints, residents in rural or remote areas, elderly populations in care homes or with mobility issues, people with disabilities, the homeless and marginalized populations, and those lacking awareness about oral health. The American Dental Association (ADA) report reveals that between 2013 and 2016, approximately 15.2% of the U.S. population required dental care but did not receive it (Gupta and Vujicic 2019). Consequently, limited access to dental care leads to inadequate periodontal examination and treatment, worsening periodontal disease, compromising oral functionality, exacerbating systemic health ailments, and imposing financial burdens on these already vulnerable groups. The impact is far-reaching, affecting not just oral health but overall well-being and quality of life (Oral Health in America 2021).

Machine learning (ML) algorithms have proven to be highly effective in handling complex tabular datasets and extracting meaningful patterns. Recently, several imaging-based machine learning models have been developed for periodontitis evaluation, almost all requiring patients’ radiographic images, such as dental X-rays or CT scans (Alotaibi et al. 2022; Jiang et al. 2022; Lee et al. 2022; Muhammed Sunnetci et al. 2022). However, individuals with limited access to dental care, as mentioned earlier, are often unable to obtain up-to-date dental radiographs. Therefore, developing a home-based, easily accessible periodontitis evaluation tool that does not rely on radiographic images is crucial for these individuals.

In this study, we validated the concept of developing an innovative data-driven, ML-based tool, which is imaging-independent, that allows individuals to conduct preliminary assessments of periodontitis conveniently at home. Our approach harnesses the power of ML and builds upon established evidence from scientific studies regarding the strong correlations between periodontitis and various demographic, lifestyle, and health factors. For instance, extensive research has unequivocally demonstrated the significant risk posed by diabetes in relation to periodontitis (Păunică et al. 2023). Similar approaches have been applied to cardiovascular disease, where ML models have demonstrated reliable accuracy in predicting death from heart disease by analyzing health and nutrition variables (Martin-Morales et al. 2023). Additionally, previous studies have well-documented the influence of demographic factors (e.g., age, gender, and race) and nutrition, which are closely linked to periodontal health, further reinforcing the credibility and reliability of our model’s predictions (Martinon et al. 2021; Selvaraj et al. 2021). By integrating these correlations, our model is capable of providing an assessment of periodontitis. Furthermore, the feature importance analysis can be used for questionnaire design, ensuring that the most influential factors are effectively captured for the real-world application of this tool (see workflow diagram in Figure 1a). Additionally, our novel approach of analyzing misclassified populations provided valuable insights for data interpretation, refining the model and uncovering complex relationships between periodontitis and its risk factors, such as menopause in females.

**Figure 1.**
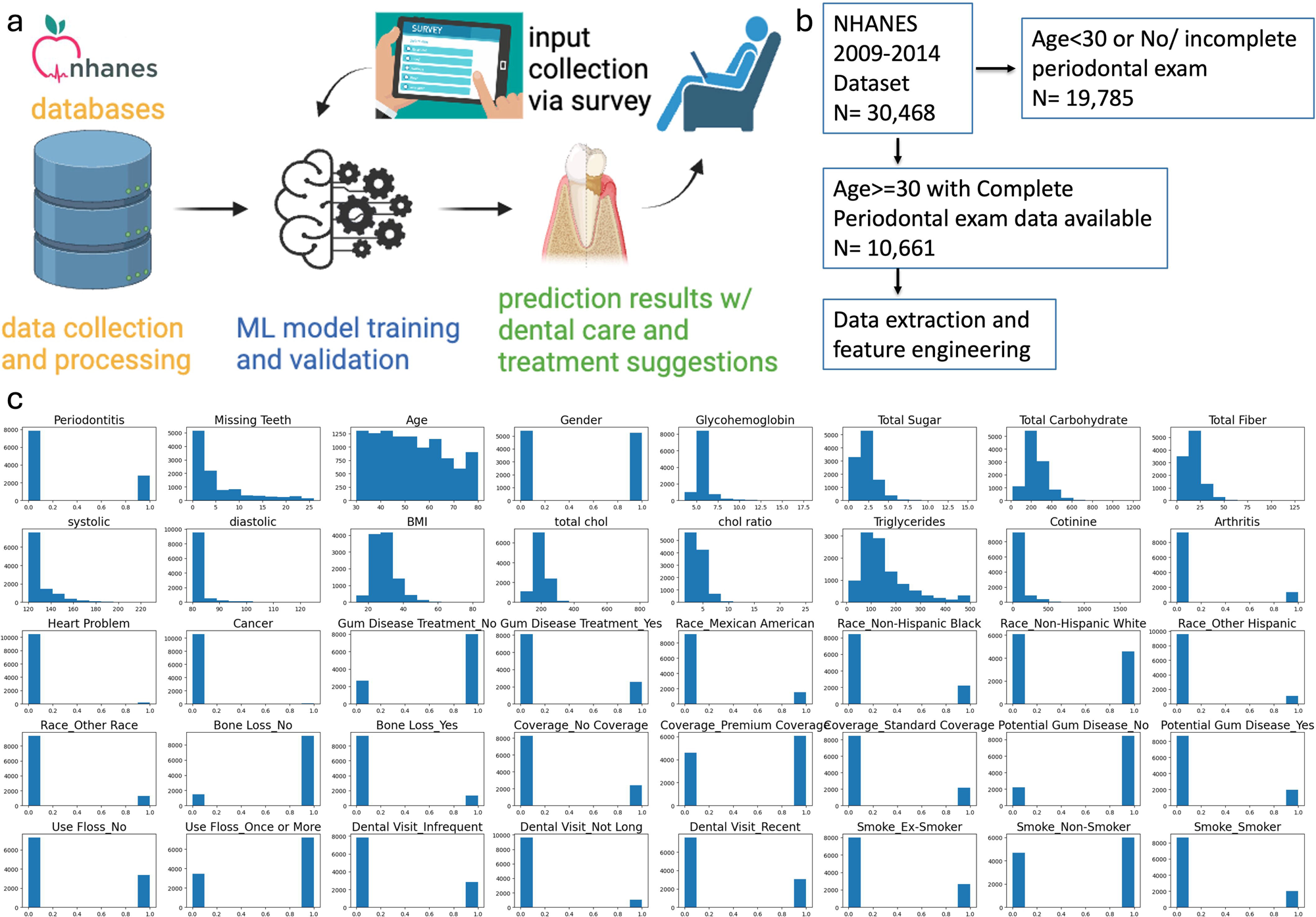
a. Workflow diagram illustrating the machine learning pipeline for home-based periodontitis assessment. b. NHANES dataset filtering process, showing the selection criteria for individuals aged 30 and above with complete periodontal exam data. c. Distribution of key features in the processed, AI-ready dataset used for model training and analysis.

## 2. Materials and Methods

### 2.1 Data Source

In this study, we utilized the ML algorithms to process the currently available datasets from 2009 to 2014, which encompasses information from over 25,000 individuals aged 30 and older, across the United States, from the National Health and Nutrition Examination Survey (NHANES) database. NHANES is a comprehensive program of studies that aims to assess the health and nutritional status of the US population. It stands out from other surveys as it combines interviews and physical examinations, providing a well-rounded set of information, including demographic specifics, nutritional profiles, data on various diseases, as well as oral health surveys and periodontal examinations. The comprehensive data allowed us to extract relevant data for periodontal health indicators and develop a machine-learning prototype specifically designed for predicting periodontitis. The study protocol was approved by the Institutional Review Board (IRB) of Tufts University (STUDY00004318).

### 2.2 Data Collection and Process

Following IRB approval, we extracted data from 2009-2014 for over 25,000 U.S. individuals aged 30 and above. This age group, characterized by mature dentitions and prolonged exposure to risk factors, is a strategic choice for periodontitis research, especially considering the higher prevalence of periodontitis in this population (Eke et al. 2015). We then thoroughly examined each variable used in this study and transformed individual variables or groups of variables into clinically meaningful features related to demographics, nutrition intake, systemic health and self-reported oral health and habits. These features, as described in the introduction section, have been previously reported with a strong correlation with periodontitis. Through meticulous feature engineering and selection, we identified 13 categorical and 14 numerical features (see Table 1).

**Table 1.** Categorical and numerical features extracted from the NHANES database.

Categorical features
| Feature Name | Classes | Feature Name | Classes |
| --- | --- | --- | --- |
| Race | Mexican American, Non-Hispanic Black, Non-Hispanic White, Other Hispanic, Other Race | May Have Gum Disease | Yes, No, Unknown |
| Sex | Male, Female | Had Gum Disease Treatment | Yes, No, Unknown |
| Insurance Coverage | No Insurance, Standard Coverage, Premium Coverage, Unknown | Been Told of Bone Loss around Teeth | Yes, No, Unknown |
| Diabetes | Diabetic w/o medication, Diabetic w/ medication, Prediabetes, Healthy, Unknown | Last Dental Visit | <6 months, 6-12 months, >12 months, Unknown |
| Smoke/Used to Smoke | Smoker, Recent-Quit-Smoker, Ex-Smoker, Infrequent-Smoker, Non-Smoker, Unknown | Has Arthritis | Yes, Unknown |
| Use Floss | No, Once or More, Unknown | Has Heart Problem | Yes, Unknown |
| Has Cancer | Yes, Unknown |  |  |

Continuous numerical features
| Feature Name | Mean (Stdev) | Feature Name | Mean (Stdev) |
| --- | --- | --- | --- |
| Number of Missing Teeth | 5.1 (6.3) | Glycohemoglobin | 5.8 (1.1) |
| Age | 52 (14) | Total Cholesterol Level | 197.6 (41.7) |
| Total Daily Sugar Intake | 2.25 (1.31) | HDL/LDL ratio | 4.04 (1.48) |
| Total Daily Carb Intake | 249 (105) | Triglycerides Level | 159.6 (144.3) |
| Total Daily Fiber Intake | 176 (9.5) | Cotinine Level | 53.8 (126.2) |
| Systolic Blood Pressure | 128.8 (13.6) | Number of cigarettes past month | 336.4 (298.6) |
| Diastolic Blood Pressure | 81.5 (4.1) | Time since quitted smoking (Years) | 18.9 (14.3) |

To assess periodontitis severity, we applied the 2017 classification procedure recommended by recommended by the American Academy of Periodontology (AAP) and European Federation of Periodontology (EFP) (Tonetti et al. 2018), while adapting steps from previous studies (Tonetti and Sanz 2019; Brito et al. 2022; Patel et al. 2022). We determined and labeled periodontitis severity for each record, using the available data on dental and periodontal examination results from NHANES. Patients are classified as healthy if they have no interproximal clinical attachment loss (IntCAL<1) in ≥2 non-adjacent teeth, and no buccal or lingual attachment loss (MidCAL) ≥3 mm with probing depth (PD) >3 mm in ≥2 teeth. Stage I periodontitis is defined by interproximal clinical attachment loss of 1 to 2 mm with a maximum probing depth ≤4 mm, Stage II by interproximal clinical attachment loss of 3 to 4 mm with a maximum probing depth ≤5 mm, and Stages III and IV by interproximal clinical attachment loss ≥5 mm with probing depth ≥6 mm. Stage III is further defined by having ≥10 occlusive pairs, while Stage IV is defined by having <10 occlusive pairs (Supplementary Figure 1). Individuals were categorized into three classes: severe cases (stage III and stage IV periodontitis) and non-severe cases (healthy, stage I and stage II periodontitis), and unknown. Excluding the “unknown” group for supervised learning, our dataset included 10,661 records with defined periodontal conditions (Figure 1b).

We further divide the dataset into training, validation, and test sets with a random split of 8:1:1 ratio, resulting in 8,529 training samples, 1,066 validation samples, and 1,066 test samples. Stratification is used to ensure the distribution of the classes in the splits mirrors the original distribution.

For the variables and features with unknown/refused to respond/missing values, we use two strategies to fill in:

- Unknown category: for categorical features, we have introduced an “unknown” class to represent non-determined values.
- Imputation: for numerical features, we have adopted imputation following best practices (Jakobsen et al. 2017).

We further transformed the extracted features with the following procedure:

- One-hot encoding on the categorical features and remove the “unknown” features from one-hot encoding as they do not provide useful information.
- Min-Max standardization on the numerical features to convert their range to [0, 1]. The min/max values are extracted only based on the train split.

With the transformation above, we have arrived at cleaned, standardized inputs with a total 40 features (13 numerical and 27 binary). The data distribution is shown in Figure 1c.

### 2.3 Model Training and Evaluation

To establish a quantitative link between the patients’ surveyable features and their periodontal examination outcome, various machine learning classifiers were employed to model the feature-label relation. We compared five commonly used machine learning algorithms using scikit-learn (Pedregosa et al.) in four categories for binary classification problems, and used the validation set for all the hyperparameter tuning:

- Logistic Regression (LR)
- Ensemble Algorithms: Random Forest (RF) and Histogram Gradient Boosting (HGBT)
- Support Vector Machine (SVM)
- Small Neural Network: 3-layer Multi-Layer Perceptron (MLP)

We first established a baseline model with Logistic Regression. With a total of 40 input features, the parameters of the model are *w_ij_* and *b_j_*, where *i* ∊ {0, …, 39} and the output logit *z*

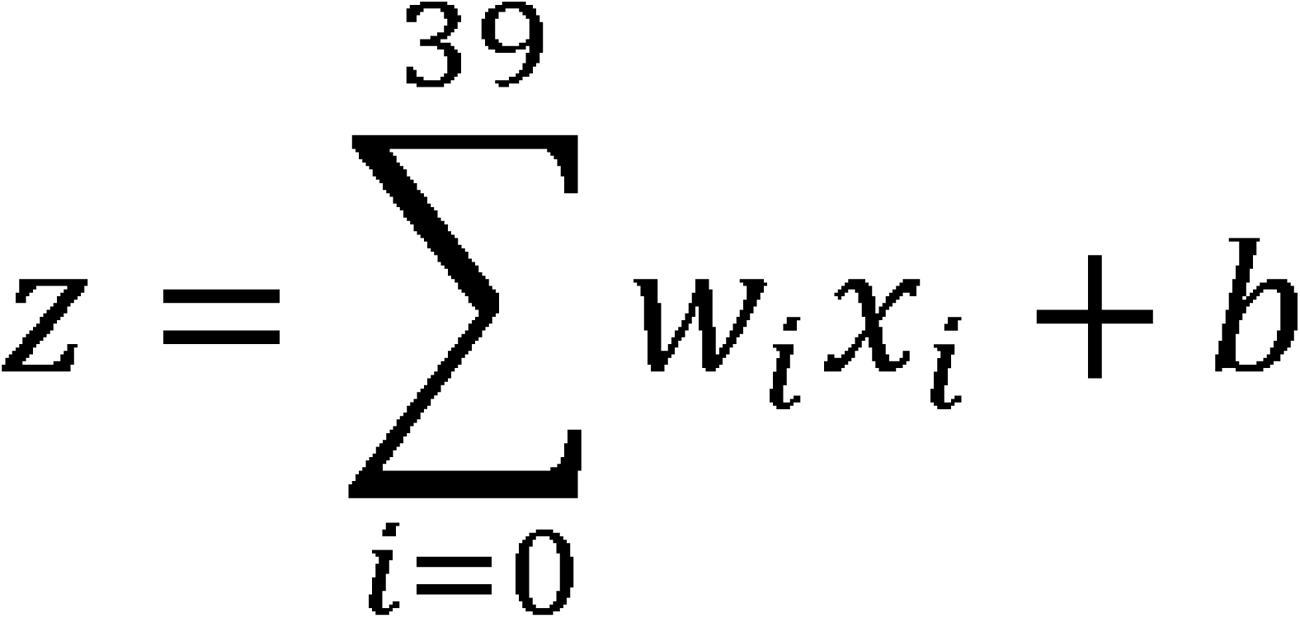

With log loss (cross-entropy), the parameters are trained to minimize

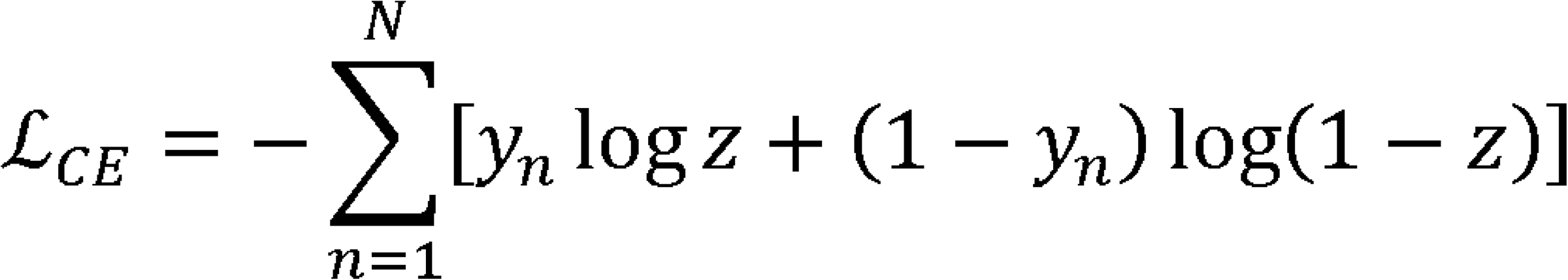

where *y_n_* is the label of the n^th^ sample (i.e., *y_n_* =1 if the n^th^ sample has periodontitis).

Since the dataset is imbalanced with the number of samples in “non-severe” and “Severe” classes, class weights are used to balance the loss from different classes such that the algorithm performs as a well-behaved maximum a posteriori (MAP) estimator.

<u>Ensemble Methods.</u> Random Forest (RF) and Histogram-Based Gradient Boosting (HGBT) are ensemble algorithms known for handling large, complex datasets. Despite potential issues with overfitting and model size, they can be effectively configured to avoid these pitfalls. In our study, we adjusted both RF and HGBT models to handle imbalanced classes and high input dimensions, using shallow trees (depth <= 10) and many samples at leaf nodes (>=30) to prevent overfitting. We also increased the number of RF estimators for smoother predictions.

<u>Support Vector Machine</u> is a versatile binary classification algorithm. It uses a high-dimensional feature space to distinguish between classes. In our study, we enhanced SVM’s robustness using a radial-basis-function (RBF) kernel, stronger regularization, and class weights.

<u>The 3-layer MLP</u> is a type of neural network that can capture complex patterns in data. In our study, we optimized the MLP by adjusting the number of neurons in each layer and the learning rate. We used ReLU activation functions for the hidden layers and a softmax activation function for the output layer. To prevent overfitting, we used a small size MLP of 32 and 8 neurons in the hidden layers, and early stopping based on validation loss. The model was trained using the Adam optimizer with a learning rate of 0.001 and a batch size of 200.

<u>Feature Reduction</u> techniques were employed to enhance model performance and interpretability by eliminating less informative features. The primary goal was to identify and retain the most relevant features that contribute significantly to the prediction of periodontitis. Two main techniques were utilized: logistic regression recursive feature reduction and feature reduction based on feature importance from the fitted random forest model. The logistic regression recursive feature reduction method iteratively removed the least significant features, as determined by their coefficients, until the optimal subset of features was identified. Alternatively, we performed feature importance analysis from the fitted random forest model to rank features based on their contribution to the model’s predictions. And we evaluated the model performance using only the top 25 features from the two methods.

<u>Prototype Model Evaluation.</u> To evaluate our trained models, we use several metrics to assess their different aspects. These includes class-weighted precision, recall, and F1-score, all averaged per-class and weighted by class support. This weighing addresses class imbalance in the dataset, providing more accurate metrics of real-world model performance. After tuning all the model hyperparameters based on the validation set, we tested the models on the held-out test set to gauge its real-world applicability. Additionally, the area under the receiver operating characteristic curve (AUROC) analysis was used to further evaluate the models’ capability to distinguish between positive and negative classes.

## 3. Results

### Model performance

We first established a baseline model, and then trained additional classifiers such as RF, HGBT and SVM. A thorough hyperparameter search yielded consistent performance across models. Figure 2a displays the metrics, including the precision, recall and F1 score of the models on training and test set. The models demonstrate strong classification performance without overfitting, which is desirable for a well-behaved model.

**Figure 2.**
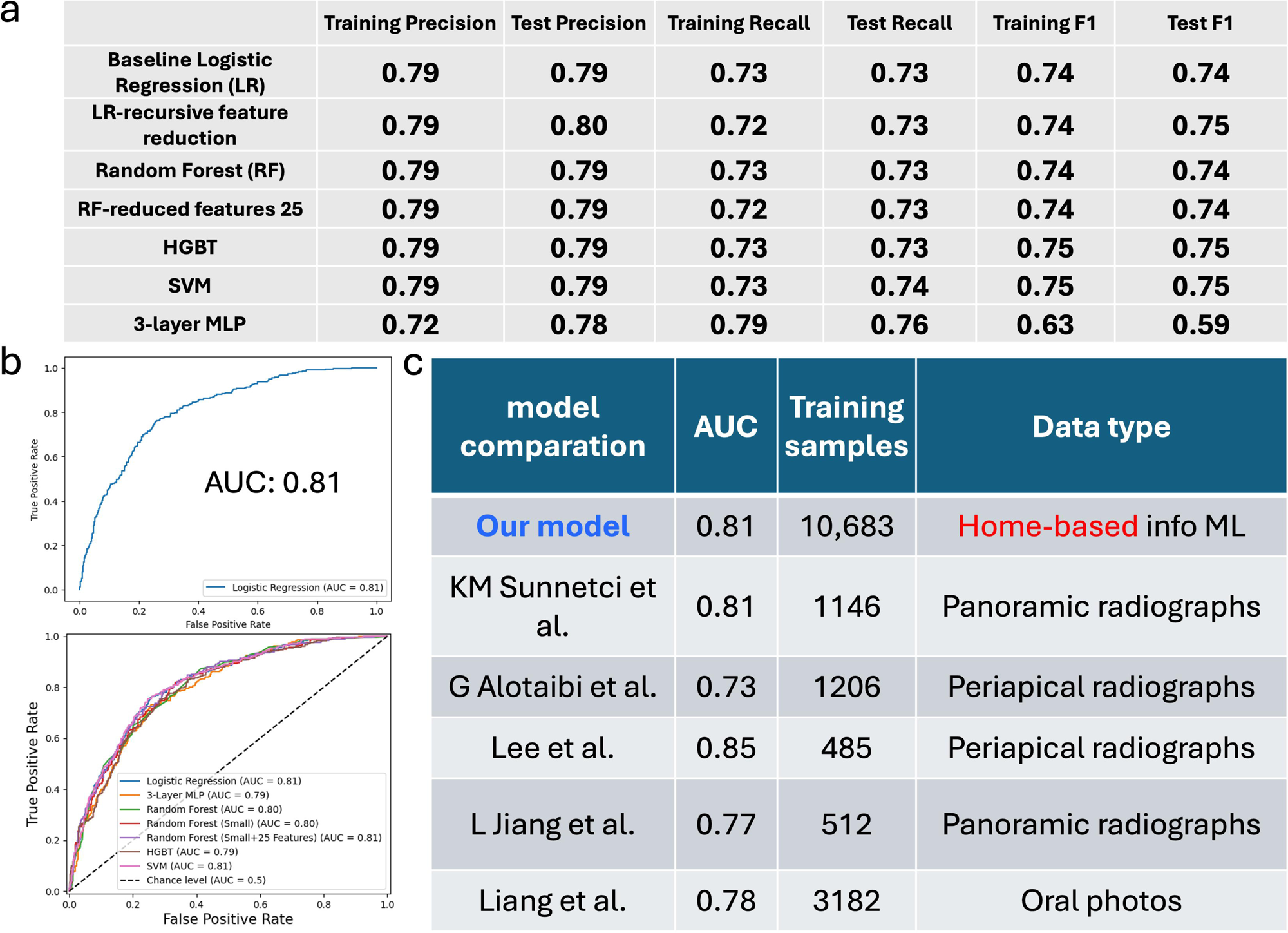
a. Comparison of different models, including Logistic Regression (LR), 3-layer Multi-Layer Perceptron (MLP), Random Forest (RF), Histogram-Based Gradient Boosting Trees (HGBT), and Support Vector Machine (SVM), based on training and test performance metrics. b. ROC curve of LR model (top) comparison of ROC curves for other models (bottom). c. Performance comparison of our model with radiographic-based models, highlighting differences in AUC, training sample size, and data type used.

We further evaluated the models’ capability to distinguish between positive and negative classes using the Receiver Operating Characteristic (ROC) curve. The area under the ROC curve (AUC) of our baseline model is 0.81, while the 3-layer MLP, RF, HGBT and SVM have similar AUC, ranging from 0.79 to 0.81 (Figure 2b). Since an AUC greater than 75% is considered clinically useful (Roumeliotis et al. 2024), our tool demonstrates strong potential for practical application in periodontal disease assessment.

Notably, our model was also compared with the published radiographic-based models (Liang et al. 2020; Alotaibi et al. 2022; Lee et al. 2022; Lee et al. 2022; Muhammed Sunnetci et al. 2022), and presents comparable performance (Figure 2c). This result demonstrates the predictive power of non-radiographic data on periodontal diseases and proves the feasibility of utilizing a home-based tool to enhance the accessibility of dental care, providing a means for timely detection and intervention, and ultimately improving patient outcomes.

### Feature coefficients

We conducted further analysis of the model’s features and identified the top 25 feature coefficients (Figure 3a). The top five features contributing to periodontitis prediction are age, cotinine level, HbA1c level, number of missing teeth, and gender. In contrast, regular dental care (every 6 months), non-Hispanic white race, and premium insurance coverage are negatively correlated with periodontitis. We are currently developing a function that can search and retrieve relevant blood test results directly from test reports or PCP notes, which patients can access through their online medical portals and upload to our tool. Additionally, the feature coefficients guide the design of our questionnaire, which will be used for input collection in our home-based tool via laptops or smartphones.

**Figure 3.**
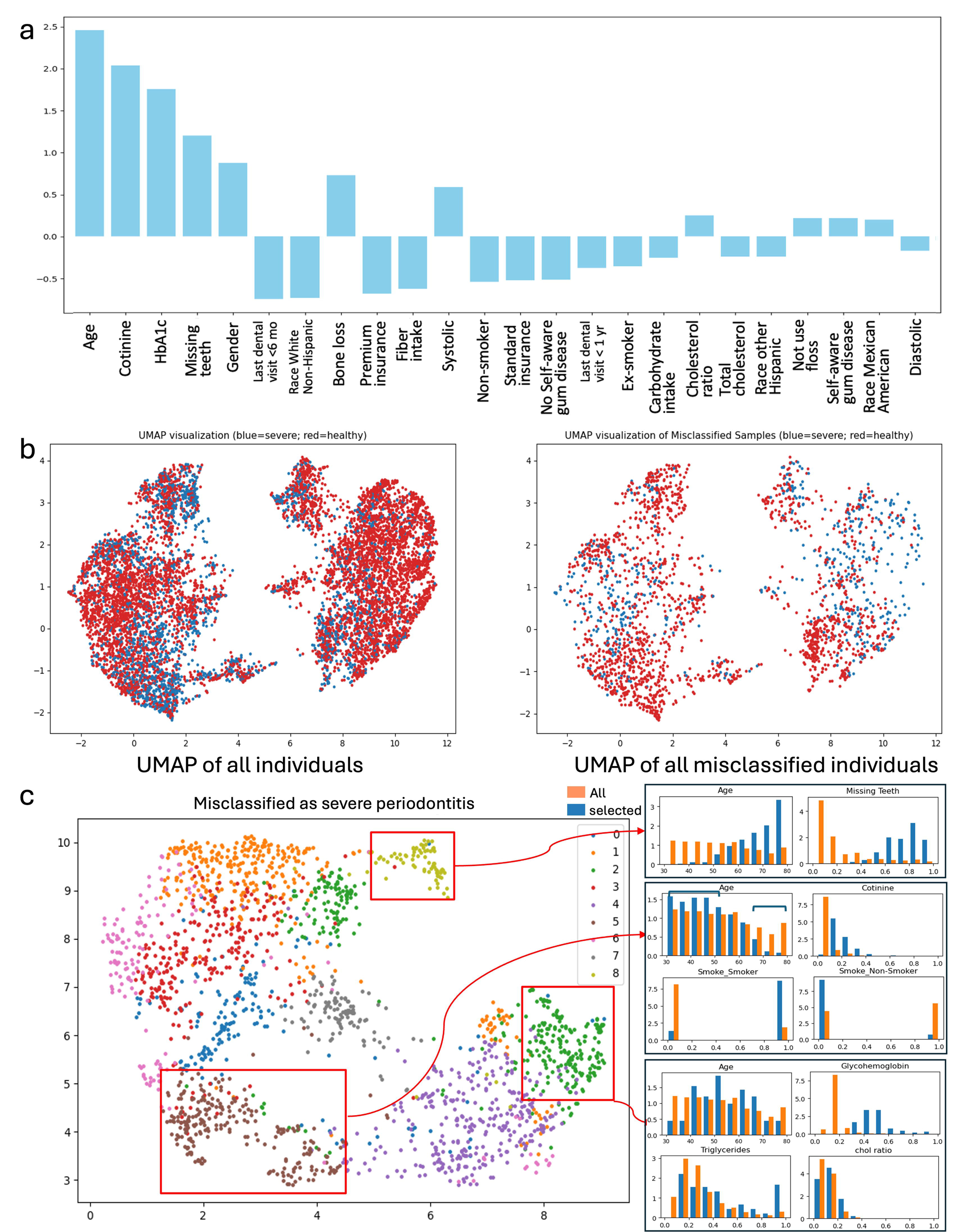
a. Feature coefficients indicating the relative importance of factors. b. UMAP visualization of all individuals (left) and misclassified individuals (right), where blue represents severe periodontitis cases and red represents healthy individuals. c. Identification of clinically meaningful subpopulations within cases misclassified as severe periodontitis, with additional demographic and clinical feature distributions shown for selected subgroups.

### Feature analysis of the misclassified samples

As dentist-scientists, we did further feature analysis and value the data interpretation, which can help identify the clinical relevance of our results and further improve the performance of our ML model. Principal Component Analysis (PCA) is a technique commonly used in bioinformatics (Ma and Dai 2011) to analyze and visualize high-dimensional data, which contains a large number of variables, such as gene expression levels or proteomic data. For the first time, we applied PCA to feature analysis and data interpretation in the periodontitis prediction model. We utilized the 25 feature coefficients, performed PCA, and visualized the results using UMAP (Uniform Manifold Approximation and Projection) and profiled the features of sub-clusters. This approach allowed us to explore the relationships among features and gain insights into the underlying patterns within the data. In Figure 3b, all individuals and misclassified samples are presented on the UMPAs, respectively.

The “misclassified as severe periodontitis” cases represent the false positive instances. We identified several sub-populations within these samples that exhibit clinically meaningful patterns. For example: 1) The cluster framed at the top in Figure 3c shows a significant concentration of older individuals with a high number of missing teeth. However, the NHANES database does not provide information on the cause of tooth loss, which may partially explain the misclassification of this sub-population. 2) The cluster on the right represents adults aged 40-65 with dyslipidemia and high HbA1c levels. While they are at high risk, they have not yet developed severe periodontitis. 3) The cluster on the left indicates that younger, smoking adults may not yet have severe periodontitis, but as they age (60-80 years), they are at a much higher risk of developing severe periodontitis compared to non-smokers.

Moreover, In the “misclassified as non-severe” (false negative) cases, we identified an intriguing subpopulation consisting exclusively of females, primarily aged 45-64 years, who are non-smokers, non-diabetic, yet have severe periodontitis that was not correctly diagnosed (Figure 4a, 4b). To further investigate, we extracted all 45-64-year-old females (n=764) from the training set (n=8,528) and found that the model’s performance on this subgroup was significantly lower compared to its performance on the entire training set (Figure 4c, 4d). This discrepancy led us to question whether menopause could be a contributing factor to misclassification in this subgroup.

**Figure 4.**
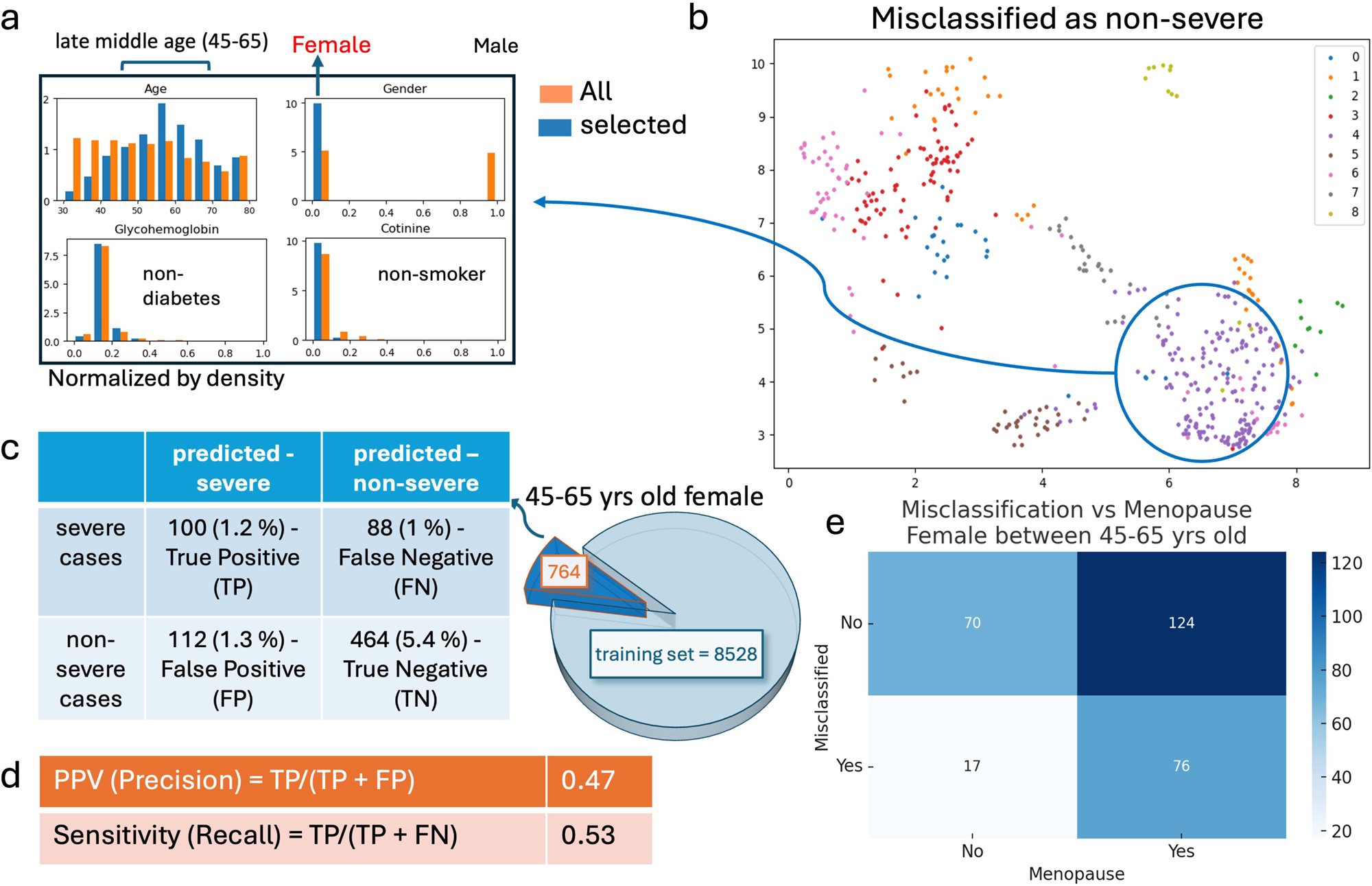
a. A subpopulation of females (mainly aged 45-64) with unique characteristics who were misclassified as non-severe cases. b. Visualization of all cases misclassified as severe periodontitis. c. Distribution of all females aged 45-65 in the training set. d. Model precision and sensitivity for this subgroup. e. Heatmap showing the correlation between misclassification and menopause.

A literature review on the association between periodontal disease and menopause revealed controversial findings (Dutt et al. 2013). To explore this relationship further, we extracted menopause-related features from the Reproductive Health (RHQ) section of NHANES and successfully identified the menopause status of 387 females aged 45-65 in our dataset. We found that 81.7% of misclassified cases in this subpopulation were menopausal individuals. Furthermore, when analyzing the relationship between misclassification and menopause (Figure 4e), we observed that misclassification was more frequent in menopausal women (38.0%) compared to non-menopausal women (19.5%), suggesting a possible positive correlation between menopause and misclassification.

This finding not only provides insights into the potential role of menopause in model misclassification but also presents an opportunity to enhance model performance by incorporating menopause-related features. Future iterations could integrate hormonal factors to improve classification accuracy, particularly for postmenopausal women and individuals undergoing hormonal therapy, thereby addressing potential biases and improving predictions for an underrepresented subgroup in machine learning models for periodontitis.

## 4. Discussion

Artificial intelligence (AI) has shown great potential in healthcare, enabling early disease detection, continuous monitoring, precision treatment planning (Khalifa and Albadawy 2024). In dentistry, AI is transforming the field by enhancing diagnostic accuracy, streamlining clinical decision-making, and improving patient management, ultimately leading to more efficient and effective dental treatments and improved patient oral health (Mahesh Batra and Reche). Our goal is to broaden the application of AI in dentistry by leveraging it to enhance access to dental care.

One of the key advantages of our model is its ability to provide a non-image-based approach to periodontitis assessment, setting it apart from existing AI models that primarily rely on radiographic data and are therefore limited to clinical use, hindering at-home applications. A recent review analyzing 12 studies highlighted that most AI models for periodontitis detection use convolutional neural networks (CNNs) trained on radiographic images (Jundaeng et al. 2025). However, these studies often suffer from small sample sizes—for example, one study included only 110 patients (Ossowska et al. 2022), limiting their generalizability. Our model, trained on a large-scale dataset, not only ensures robustness but also eliminates the need for radiographic imaging, enabling home-based self-assessment of periodontitis. This innovation democratizes dental healthcare by empowering individuals—especially those unaware of their undiagnosed periodontal conditions—to take proactive steps in managing their oral health.

Furthermore, radiographic findings primarily reflect the outcome of periodontitis, while our model investigates underlying risk factors. By exploring the relationships between periodontitis and demographic, systemic health, and lifestyle influences, our model facilitates personalized risk assessment and prediction. This approach allows for early intervention and patient-centered dental solutions, a concept reinforced by prior research linking systemic factors with periodontal disease (Pitchika et al. 2024).

On the other hand, unlike previous basic science and clinical studies that focus on understanding the pathophysiology of periodontitis in systemic disorders (Jain et al. 2021; Wu et al. 2022) or identifying the risk indicator of periodontitis (Eke et al. 2016), our model builds upon these well-established risk factors to provide a predictive approach to periodontitis. This novel and practical application transforms foundational research into a valuable tool for preventive dentistry, facilitating early intervention and patient-centered dental solution.

Beyond model development, our study emphasizes data interpretation. By visualizing severe and non-severe periodontitis, along with misclassified cases using UMAP, we extracted clinically meaningful subpopulations. One notable finding was the misclassification of periodontitis severity in postmenopausal women. The relationship between menopause and periodontal disease remains controversial, with studies presenting conflicting evidence.

Some studies suggest a positive correlation, indicating that hormonal changes during menopause contribute to periodontal disease progression. For example, For example, a study using data from the Rafsanjan Cohort Study found that hormonal changes during menopause negatively impacted periodontal health, leading to CAL and PD (Rafiei et al. 2022). Similarly, research on early menopause in Korean postmenopausal women revealed a significant association between menopause and periodontitis (Lee et al. 2018). On the other hand, some studies report no significant association. A cross-sectional study in a Portuguese population found only a slight increase in attachment loss among menopausal women, with the difference not reaching statistical significance (Alves et al. 2015). Another study concluded that while periodontal disease and bone loss share common risk factors, periodontitis itself may not be a direct consequence of menopause, questioning the direct impact of hormonal changes (Lee 2022).

These inconsistencies suggest that menopause’s impact on periodontal health is likely influenced by additional factors such as genetics, lifestyle, and overall systemic health. Our data suggests that menopause may contribute to misclassification in AI predictions, highlighting the need for further research to refine our model.

While our model demonstrates strong performance, its predictive accuracy is constrained by limitations in the dataset itself rather than by the machine learning algorithms. Several factors contribute to misclassification: a. Data Quality: NHANES periodontal examination protocols have been reported to misclassify periodontitis cases, affecting validity for research and surveillance (Eke et al. 2010). b. Sample Size: Machine learning models require large-scale datasets to ensure optimal training. Our dataset originally categorized periodontitis into five stages (healthy, stage 1, stage 2, stage 3, stage 4) based on the 2017 classification system.

However, the healthy class contained only 360 samples in the training set (Supplementary Table 1), leading to class imbalance and impacting model performance. c. Feature Availability: NHANES data provides valuable demographic and health information but lacks longitudinal patient follow-up, limiting our ability to assess disease progression over time.

To overcome data limitations and enhance model performance, we plan to integrate larger, high-quality electronic dental records (EDR). Specifically, we plan to utilize AxiUm data from Tufts University School of Dental Medicine (TUSDM), which includes over 150,000 patient records, significantly surpassing the 25,000 records in NHANES. High-quality EDR data has demonstrated superior predictive capabilities in periodontitis classification, as seen in a recent study at Temple University, where they used EDR data from Temple Dental School to predict periodontitis staging using CAL and PD with 100% sensitivity, specificity, and accuracy due to the excellent data quality (Patel et al. 2024). Additionally, this database provides time-series data, which will allow us to incorporate longitudinal modeling for periodontitis grading and progression prediction. By applying advanced time-series ML models, such as Long Short-Term Memory networks (LSTMs) and transformers, we can refine our model to better track periodontitis progression across different patient populations.

In the long term, we aim to deploy our AI-driven tool as a user-friendly online questionnaire, providing a practical and accessible solution for preliminary periodontal assessments. This tool will bridge the gap in early detection for individuals with limited access to dental care. As data accumulates, incorporating longitudinal insights will further improve prediction accuracy and expand the tool’s clinical utility. Additionally, leveraging time-series models will allow us to better predict disease progression and patient-specific risk levels, ensuring a more dynamic and adaptable AI-driven system.

## Conclusion

Our machine-learning model has demonstrated robust predictive performance, establishing a solid foundation for future enhancements. With continued refinement, we anticipate achieving even greater accuracy in predicting periodontitis. This tool has the potential to address critical gaps in dental healthcare accessibility, benefiting individuals who may already have undiagnosed periodontal disease requiring early intervention. By leveraging AI, our approach contributes to advancing preventive dentistry and improving oral health outcomes, ultimately enhancing overall well-being and quality of life for a broader population.

## Data Availability

All results of this study are described in the manuscript.

## Acknowledgments

The author(s) would like to acknowledge the support by the National Institutes of Health (NIH) grants R01 DE30074 to Jake Chen.

---

**Supplementary Figure 1.**
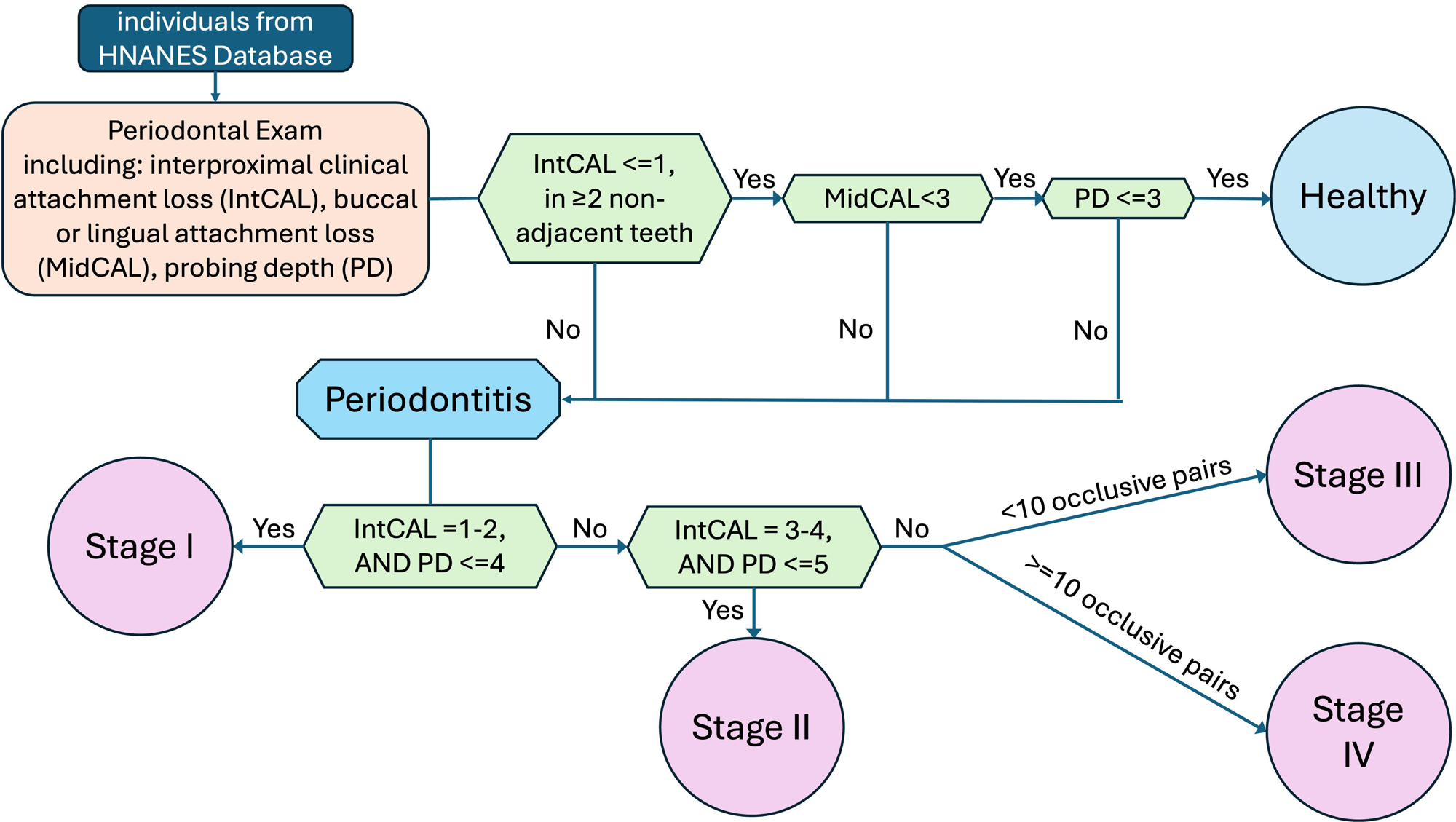
Diagnosis steps for periodontitis staging on NHANES data.

**Supplementary Table 1.** The train, validation and test sets.

|  | All individuals | Healthy | Stage I | Stage II | Stage III | Stage IV |
| --- | --- | --- | --- | --- | --- | --- |
| Total | 10661 | 450 | 1323 | 6074 | 1239 | 1575 |
| Training set | 8529 | 360 | 1058 | 4859 | 991 | 1260 |
| Validation set | 1066 | 45 | 132 | 607 | 124 | 158 |
| Test set | 1066 | 45 | 132 | 607 | 124 | 158 |

## Notes

### Competing Interest Statement

The authors have declared no competing interest.

### Author Declarations

The source data is openly available from the NHANES before the initiation of this study. The study protocol was approved by the Institutional Review Board (IRB) of Tufts University (STUDY00004318).

## Reference

Abusleme L, Hoare A, Hong B-Y, Diaz PI. 2021. Microbial signatures of health, gingivitis, and periodontitis. Periodontol 2000. 86(1):57–78.

Alotaibi G, Awawdeh M, Farook FF, Aljohani M, Aldhafiri RM, Aldhoayan M. 2022. Artificial intelligence (AI) diagnostic tools: utilizing a convolutional neural network (CNN) to assess periodontal bone level radiographically—a retrospective study. BMC Oral Health. 22(1):399.

Alves RC, Félix SA, Rodriguez-Archilla A, Oliveira P, Brito J, Dos Santos JM. 2015. Relationship between menopause and periodontal disease: a cross-sectional study in a Portuguese population. Int J Clin Exp Med. 8(7):11412–11419.

Brito LF, Taboza ZA, Silveira VR, Teixeira AK, Rego RO. 2022. Diagnostic accuracy of severe periodontitis case definitions: Comparison of the CDC/AAP, EFP/AAP, and CPI criteria. Journal of Periodontology. 93(6):867–876.

Chapple ILC, Genco R, Workshop* on behalf of working group 2 of the joint E. 2013. Diabetes and periodontal diseases: consensus report of the Joint EFP/AAP Workshop on Periodontitis and Systemic Diseases. Journal of Periodontology. 84(4S):S106–S112.

Dietrich T, Sharma P, Walter C, Weston P, Beck J. 2013. The epidemiological evidence behind the association between periodontitis and incident atherosclerotic cardiovascular disease. Journal of Periodontology. 84(4S):S70–S84.

Dutt P, Chaudhary S, Kumar P. 2013. Oral Health and Menopause: A Comprehensive Review on Current Knowledge and Associated Dental Management. Ann Med Health Sci Res. 3(3):320– 323.

Eke PI, Borgnakke WS, Genco RJ. 2020. Recent epidemiologic trends in periodontitis in the USA. Periodontol 2000. 82(1):257–267.

Eke PI, Dye BA, Wei L, Slade GD, Thornton-Evans GO, Borgnakke WS, Taylor GW, Page RC, Beck JD, Genco RJ. 2015. Update on Prevalence of Periodontitis in Adults in the United States: NHANES 2009 to 2012. J Periodontol. 86(5):611–622.

Eke PI, Thornton-Evans GO, Wei L, Borgnakke WS, Dye BA. 2010. Accuracy of NHANES periodontal examination protocols. J Dent Res. 89(11):1208–1213.

Eke PI, Wei L, Thornton-Evans GO, Borrell LN, Borgnakke WS, Dye B, Genco RJ. 2016. Risk Indicators for Periodontitis in US Adults: NHANES 2009 to 2012. Journal of Periodontology. 87(10):1174–1185.

Gupta N, Vujicic M. 2019. Main Barriers to Getting Needed Dental Care All Relate to Affordability. Health Policy Institute Research Brief American Dental Association.

Jain P, Hassan N, Khatoon K, Mirza MohdA, Naseef PP, Kuruniyan MS, Iqbal Z. 2021. Periodontitis and Systemic Disorder—An Overview of Relation and Novel Treatment Modalities. Pharmaceutics. 13(8):1175.

Jakobsen JC, Gluud C, Wetterslev J, Winkel P. 2017. When and how should multiple imputation be used for handling missing data in randomised clinical trials – a practical guide with flowcharts. BMC Medical Research Methodology. 17(1):162.

Jiang L, Chen D, Cao Z, Wu F, Zhu H, Zhu F. 2022. A two-stage deep learning architecture for radiographic staging of periodontal bone loss. BMC Oral Health. 22(1):106.

Jundaeng J, Chamchong R, Nithikathkul C. 2025. Periodontitis diagnosis: A review of current and future trends in artificial intelligence. Technol Health Care. 33(1):473–484.

Khalifa M, Albadawy M. 2024. Artificial Intelligence for Clinical Prediction: Exploring Key Domains and Essential Functions. Computer Methods and Programs in Biomedicine Update. 5:100148.

Larvin H, Gao C, Kang J, Aggarwal VR, Pavitt S, Wu J. 2023. The impact of study factors in the association of periodontal disease and cognitive disorders: systematic review and meta-analysis. Age and Ageing. 52(2):afad015.

Lee C-T, Kabir T, Nelson J, Sheng S, Meng H-W, Van Dyke TE, Walji MF, Jiang X, Shams S. 2022. Use of the deep learning approach to measure alveolar bone level. J Clin Periodontol. 49(3):260–269.

Lee Y. 2022. Association between osteoporosis and periodontal disease among menopausal women: The 2013–2015 Korea National Health and Nutrition Examination Survey. PLoS One. 17(3):e0265631.

Lee Y-H, Kim S-M, Ahn E. 2018. Relationship between Early Menopause and Periodontal Disease in Korean Postmenopausal Women. Journal of Dental Hygiene Science. 18(5):312– 318.

Liang Y, Fan HW, Fang Z, Miao L, Li W, Zhang X, Sun W, Wang K, He L, Chen X “Anthony.” 2020. OralCam: Enabling Self-Examination and Awareness of Oral Health Using a Smartphone Camera. In: Proceedings of the 2020 CHI Conference on Human Factors in Computing Systems. New York, NY, USA: Association for Computing Machinery. (CHI ’20). p. 1–13. [accessed 2025 Feb 18]. https://dl.acm.org/doi/10.1145/3313831.3376238.

Liccardo D, Cannavo A, Spagnuolo G, Ferrara N, Cittadini A, Rengo C, Rengo G. 2019. Periodontal Disease: A Risk Factor for Diabetes and Cardiovascular Disease. Int J Mol Sci. 20(6):1414.

Ma S, Dai Y. 2011. Principal component analysis based methods in bioinformatics studies. Briefings in Bioinformatics. 12(6):714–722.

Mahesh Batra A, Reche A. A New Era of Dental Care: Harnessing Artificial Intelligence for Better Diagnosis and Treatment. Cureus. 15(11):e49319.

Martin-Morales A, Yamamoto M, Inoue M, Vu T, Dawadi R, Araki M. 2023. Predicting Cardiovascular Disease Mortality: Leveraging Machine Learning for Comprehensive Assessment of Health and Nutrition Variables. Nutrients. 15(18). [accessed 2023 Dec 20]. https://www.ncbi.nlm.nih.gov/pmc/articles/PMC10534618/.

Martinon P, Fraticelli L, Giboreau A, Dussart C, Bourgeois D, Carrouel F. 2021. Nutrition as a Key Modifiable Factor for Periodontitis and Main Chronic Diseases. JCM. 10(2):197.

Muhammed Sunnetci K, Ulukaya S, Alkan A. 2022. Periodontal bone loss detection based on hybrid deep learning and machine learning models with a user-friendly application. Biomedical Signal Processing and Control. 77:103844.

Oral Health in America: Advances and Challenges: Executive Summary. 2021. Bethesda (MD): National Institute of Dental and Craniofacial Research (US). [accessed 2023 Dec 12]. http://www.ncbi.nlm.nih.gov/books/NBK576536/.

Ossowska A, Kusiak A, Świetlik D. 2022. Evaluation of the Progression of Periodontitis with the Use of Neural Networks. Journal of Clinical Medicine. 11(16):4667.

Patel J, Shin D, Willis L, Zai A, Thyvalikakath T. 2024. Feasibility of Utilizing Electronic Dental Record Data and Periodontitis Case Definition to Automate Diagnosis. In: MEDINFO 2023 — The Future Is Accessible. IOS Press. p. 214–218. [accessed 2025 Feb 24]. https://ebooks.iospress.nl/doi/10.3233/SHTI230958.

Patel JS, Brandon R, Tellez M, Albandar JM, Rao R, Krois J, Wu H. 2022. Developing Automated Computer Algorithms to Phenotype Periodontal Disease Diagnoses in Electronic Dental Records. Methods of Information in Medicine. 61:e125–e133.

Păunică I, Giurgiu M, Dumitriu AS, Păunică S, Pantea Stoian AM, Martu M-A, Serafinceanu C. 2023. The Bidirectional Relationship between Periodontal Disease and Diabetes Mellitus—A Review. Diagnostics. 13(4):681.

Pedregosa F, Varoquaux G, Gramfort A, Michel V, Thirion B, Grisel O, Blondel M, Prettenhofer P, Weiss R, Dubourg V, et al. Scikit-learn: Machine Learning in Python. MACHINE LEARNING IN PYTHON.

Pitchika V, Büttner M, Schwendicke F. 2024. Artificial intelligence and personalized diagnostics in periodontology: A narrative review. Periodontology 2000. 95(1):220–231.

Rafiei M, Salarisedigh S, Khalili P, Jamali Z, Sardari F. 2022. Hormonal Fluctuations and Periodontal Status in Postmenopausal Women. International Journal of Dentistry. 2022(1):9990451.

Relationships between Demographic Factors and Chronic Conditions with Disease Severities - PMC. [accessed 2023 Dec 20]. https://www.ncbi.nlm.nih.gov/pmc/articles/PMC8583414/.

Roumeliotis S, Schurgers J, Tsalikakis DG, D’Arrigo G, Gori M, Pitino A, Leonardis D, Tripepi G, Liakopoulos V. 2024. ROC curve analysis: a useful statistic multi-tool in the research of nephrology. Int Urol Nephrol. 56(8):2651–2658.

Selvaraj S, Naing NN, Wan-Arfah N, de Abreu MHNG. 2021. Demographic and Habitual Factors of Periodontal Disease among South Indian Adults. Int J Environ Res Public Health. 18(15):7910.

Sim S-J, Kim H-D, Moon J-Y, Zavras AI, Zdanowicz J, Jang S-J, Jin B-H, Bae K-H, Paik D-I, Douglass CW. 2008. Periodontitis and the risk for non-fatal stroke in Korean adults. J Periodontol. 79(9):1652–1658.

Tonetti MS, Greenwell H, Kornman KS. 2018. Staging and grading of periodontitis: Framework and proposal of a new classification and case definition. Journal of Periodontology. 89(S1):S159–S172.

Tonetti MS, Sanz M. 2019. Implementation of the new classification of periodontal diseases: Decision-making algorithms for clinical practice and education. Journal of Clinical Periodontology. 46(4):398–405.

Wu H, Qiu W, Zhu X, Li X, Xie Z, Carreras I, Dedeoglu A, Van Dyke T, Han YW, Karimbux N, et al. 2022. The Periodontal Pathogen Fusobacterium nucleatum Exacerbates Alzheimer’s Pathogenesis via Specific Pathways. Front Aging Neurosci. 14:912709.

